# Development of Parkinson’s disease and its relationship with incidentally-discovered white matter disease and covert brain infarction in a real world cohort

**DOI:** 10.1101/2022.02.18.22271187

**Authors:** David M. Kent, Lester Y. Leung, Eric J. Puttock, Andy Y. Wang, Patrick H. Luetmer, David F. Kallmes, Jason Nelson, Sunyang Fu, Chengyi Zheng, Hongfang Liu, Alastair J. Noyce, Wansu Chen

## Abstract

**Importance:** While a link between cerebrovascular disease and cerebrovascular risk factors and subsequent development of Parkinson’s disease has been suggested, the association between covert cerebrovascular disease and subsequent Parkinson’s disease has not been rigorously examined.

**Objective:** To examine the relationship between covert cerebrovascular disease, comprised of covert brain infarction and white matter disease, discovered incidentally in routine care and subsequent Parkinson’s disease.

**Design:** Enrollment in this observational cohort study occurred between 2009 and 2019.

**Setting:** Kaiser Permanente Southern California health system.

**Participants:** Patients were ≥50 years old and received neuroimaging for a non-stroke indication.

**Exposure:** Incidental covert brain infarction and white matter disease identified by natural language processing of neuroimaging reports. Natural language processing also classified white matter disease severity.

**Main Outcomes:** Parkinson’s disease.

**Results:** 230,062 patients were included with a total of 980,772 person-years of follow-up and a median follow-up time of 3.72 years. 1,941 cases of Parkinson’s disease were identified, with a median time-to-event of 2.35 (IQR 0.90 to 4.58) years. Covert cerebrovascular disease was identified in 70,592 (30.68%) patients, 10,622 (4.62%) with covert brain infarction and 65,814 (28.61%) with white matter disease. After adjustment for known risk factors, white matter disease was found to be associated with Parkinson’s disease, with an adjusted hazard ratio of 1.67 (1.44, 1.93) for patients <70 years and 1.33 (1.18, 1.50) for those ≥70 years. Greater severity of white matter disease was associated with an increased incidence of Parkinson’s disease per 1000 person-years, from 1.52 (1.43, 1.61) in patients without white matter disease to 4.90 (3.86, 6.13) in those with severe white matter disease. Findings were robust when more specific definitions of Parkinson’s disease were used. Covert brain infarction was not associated with Parkinson’s disease (adjusted hazard ratio=1.05 [0.88, 1.24]).

**Conclusions and Relevance:** White matter disease was associated with subsequent Parkinson’s disease, an association strengthened with younger age and increased white matter disease severity. Covert brain infarction did not appear to be associated with subsequent Parkinson’s disease.

## INTRODUCTION

Parkinson’s disease (PD) is a debilitating, insidious, and complex neurodegenerative disease that afflicts more than six million people globally, and may double in prevalence by 2040.^1^ There are no neuroprotection strategies that have been shown to decrease the risk of PD.^2^ Research is needed on PD risk factors both to identify high risk patients that might benefit from neuroprotective strategies and also to understand potentially modifiable risk factors that might be targeted to slow or prevent PD development.^3^ Emerging evidence suggests that cerebrovascular risk factors, as well as stroke itself, may be linked to subsequent development of PD.^4^

There is increasing interest in covert cerebrovascular disease (CCD) as a cerebrovascular risk factor and even as a stroke equivalent.^5^ Comprising covert brain infarction (CBI) and white matter disease (WMD), CCD is often discovered incidentally on patients who receive neuroimaging during routine clinical care. Previous studies have examined whether WMD in patients with existing PD correlates with severity or progression.^6-11^ One systematic review and meta-analysis of case-control studies suggested that white matter hyperintensities were not more prevalent or severe in patients with PD.^12^ However, case control studies are fundamentally limited because associations may be obscured through reverse causation (i.e., PD and its treatment may influence CCD). In part because the incidence of PD is relatively low, and because CCD is not identifiable using databases from routine care, no study has yet examined the association of CCD with future incidence of PD.

In previous work, we developed and validated natural language processing (NLP) algorithms to identify the presence of CCD and severity of WMD using neuroimaging reports and deployed these algorithms in a large integrated health system,^13^ thereby permitting us to examine associations between exposures and rare outcomes in a cohort by using ‘big data’. We previously found that CCD is strongly predictive of subsequent stroke^14^ and dementia,^15^ particularly in younger patients and in those with more severe WMD. In this study, we examined the association of CBI and WMD with a subsequent diagnosis of PD, using NLP of routinely obtained neuroimaging reports to identify these lesions. We also examined whether increasing severity of WMD, also ascertained through NLP of neuroimaging reports, strengthened this association.

## METHODS

### Environment

In this retrospective cohort study, we utilized health plan enrollees of Kaiser Permanente Southern California (KPSC), an integrated health care organization that serves 4.8 million individuals (approximately 19% of the region’s population) at 15 hospitals and 230+ medical offices. Enrollees’ demographics and socioeconomic status are representative of the residents residing in the region.^16^ Implemented between 2004 and 2008 at KPSC, HealthConnect®, KP’s comprehensive electronic health record (EHR), is one of the largest EHR systems in the world. This system integrates all aspects of care, including inpatient, emergency department, outpatient, pharmacy, and lab services, as well as billing and claims. All structured data used in the current study were captured from KPSC Research Data Warehouse. The study protocol was approved by Tufts Medical Center’s and KPSC’s Institutional Review Boards.

### Population

We included individuals age ≥ 50 years enrolled in the KPSC health system who received head neuroimaging (CT, MRI) for a non-stroke indication between 2009 and 2019. Exclusion criteria included prior history of PD (using codes for ICD-9 (332.0) and ICD-10 (G20)) history of ischemic stroke, dementia/Alzheimer’s disease, transient ischemic attack, or a “high probability” stroke symptom (defined in this study as aphasia, hemiparesis [including face, arm, and/or leg weakness], hemisensory loss, hemiataxia, hemineglect, visual disturbance [vision loss, diplopia], dysarthria, and dysphagia). Corresponding ICD diagnosis and ICD/CPT procedure codes are in Supplement A1, eTable 1. We additionally excluded patients with a visit reason or scan indication suggestive of cognitive symptoms or decline (e.g., confusion, disorientation, altered mental status, or dementia evaluation). Supplement A1, eTable 1 includes the complete ICD-10/CPT codes used to exclude patients.

**Table 1.** Patient demographic and clinical characteristics at baseline (n=230,062), n (%) unless otherwise stated.

| Patient characteristics | Entire cohort | Subset: patients with CBI | Subset: patients with WMD | Subset: patients without CCD<sup>a</sup> |
| --- | --- | --- | --- | --- |
| **N** | 230062 | 10622 | 65814 | 159470 |
| **Demographics** |  |  |  |  |
| **Age, mean (SD)** | 64.8 (10.37) | 72.2 (10.76) | 70.6 (10.75) | 62.2 (9.14) |
| **Female** | 142171 (61.80%) | 6130 (57.71%) | 40588 (61.67%) | 98813 (61.96%) |
| **Race/ ethnicity** |  |  |  |  |
| **Asian and Pacific Islander** | 27267 (11.85%) | 1186 (11.17%) | 7642 (11.61%) | 19089 (11.97%) |
| **African American** | 26186 (11.38%) | 1547 (14.56%) | 7376 (11.21%) | 18100 (11.35%) |
| **Hispanic** | 74922 (32.57%) | 2710 (25.51%) | 16512 (25.09%) | 57007 (35.75%) |
| **Multiple/Other/Unknown** | 3732 (1.62%) | 124 (1.17%) | 901 (1.37%) | 2774 (1.74%) |
| **Non-Hispanic white** | 97955 (42.58%) | 5055 (47.59%) | 33383 (50.72%) | 62500 (39.19%) |
| **Stroke risk factors** |  |  |  |  |
| **Atrial fibrillation (AF)** | 13107 (5.70%) | 1202 (11.32%) | 5875 (8.93%) | 6817 (4.27%) |
| **Carotid atherosclerosis** | 2186 (0.95%) | 192 (1.81%) | 1047 (1.59%) | 1077 (0.68%) |
| **Congestive heart failure (CHF)** | 10489 (4.56%) | 1105 (10.40%) | 4730 (7.19%) | 5361 (3.36%) |
| **Coronary artery disease (CAD)** | 24267 (10.55%) | 2000 (18.83%) | 9563 (14.53%) | 13951 (8.75%) |
| **Diabetes mellitus (DM)** | 59479 (25.85%) | 3716 (34.98%) | 19125 (29.06%) | 38778 (24.32%) |
| **Hypercholesterolemia (HC)** | 156591 (68.06%) | 8006 (75.37%) | 48119 (73.11%) | 104963 (65.82%) |
| **Hypertension (HTN)** | 138566 (60.23%) | 8380 (78.89%) | 46761 (71.05%) | 88285 (55.36%) |
| **Peripheral arterial disease (PAD)** | 9887 (4.30%) | 993 (9.35%) | 4402 (6.69%) | 5153 (3.23%) |
| **Tobacco use (ever)** |  |  |  |  |
| **Yes** | 105621 (45.91%) | 5553 (52.28%) | 32856 (49.92%) | 70334 (44.10%) |
| **No** | 123716 (53.78%) | 5035 (47.40%) | 32832 (49.89%) | 88561 (55.53%) |
| **Unknown** | 725 (0.32%) | 34 (0.32%) | 126 (0.19%) | 575 (0.36%) |
| **Number of stroke risk factors** | 2.3 (1.47) | 2.9 (1.58) | 2.6 (1.51) | 2.1 (1.41) |
| **Systolic blood pressure, mean (SD)** | 129.4 (13.15) | 132.8 (13.84) | 131.5 (13.05) | 128.4 (13.06) |
| **Modality** |  |  |  |  |
| **CT** | 170938 (74.30%) | 8516 (80.17%) | 34809 (52.89%) | 131822 (82.66%) |
| **MRI** | 59124 (25.70%) | 2106 (19.83%) | 31005 (47.11%) | 27648 (17.34%) |
| Antiplatelet use | 10399 (4.52%) | 704 (6.63%) | 3671 (5.58%) | 6452 (4.05%) |
| Statin use | 98996 (43.03%) | 5673 (53.41%) | 33060 (50.23%) | 63511 (39.83%) |
| Depression | 44733 (19.44%) | 2048 (19.28%) | 12833 (19.50%) | 30932 (19.40%) |
| Exercise (hour/per week), mean (SD) | 1.8 (2.54) | 1.5 (2.42) | 1.7 (2.50) | 1.8 (2.55) |
| BMI (kg/m<sup>2</sup>), mean (SD) | 28.7 (6.04) | 27.9 (5.92) | 27.9 (5.82) | 29.1 (6.09) |
| Covert brain infarction | 10622 (4.62%) | NA | 5844 (8.88%) | NA |
| White matter disease | 65814 (28.61%) | 5844 (55.02%) | NA | NA |
| Covert cerebrovascular disease<sup>a</sup> | 70592 (30.68%) | NA | NA | NA |

<sup>a</sup>Covert brain infarction or white matter disease.
BMI indicates body mass index, CBI, covert brain infarction; CCD, covert cerebrovascular disease; SD, standard deviation; WMD, white matter disease.

If there were multiple neuroimaging studies for the same individual, the first study was considered the index scan. For neuroimaging evidence of cerebral infarction to be considered “covert,” individuals were only included in the study if they did not acquire a new ICD code for a diagnosis of PD, stroke, or dementia within 180 days after the index scan. Patients who were not actively enrolled in the KPSC health plan on the index date or not continuously enrolled in the prior 12 months were also excluded (a gap of ≤45 days was allowed).

### Identification of patients with CCD

An NLP algorithm designed at Mayo Clinic and Tufts Medical Center was applied to neuroimaging reports associated with these index scans to identify individuals with documented CBI or WMD.^13^ As described in prior work, these algorithms adopted the open-source NLP pipeline MedTagger for generic NLP processing and task-specific knowledge engineering (coding of specific words and phrases referencing CBIs and WMD) to yield identification of CBI and WMD that was on par with human readers of the neuroimaging reports.^13^ This NLP algorithm achieved F-scores of 0.86 and 0.89 in CBI and WMD on 490 reports from KPSC when it was initially implemented. After retraining on those reports, the F-scores were enhanced to 0.93 and 0.92, respectively, on a separate test dataset (n=490).

We additionally used NLP to classify patients according to WMD severity into three grades: mild, moderate and severe. The development of the NLP algorithm to classify patients into different WMD classes is discussed in Supplement A2 and Supplement A2, eTable 1 and 2. Scan reports with insufficient information on severity were classified as “undetermined.”

**Table 2.**
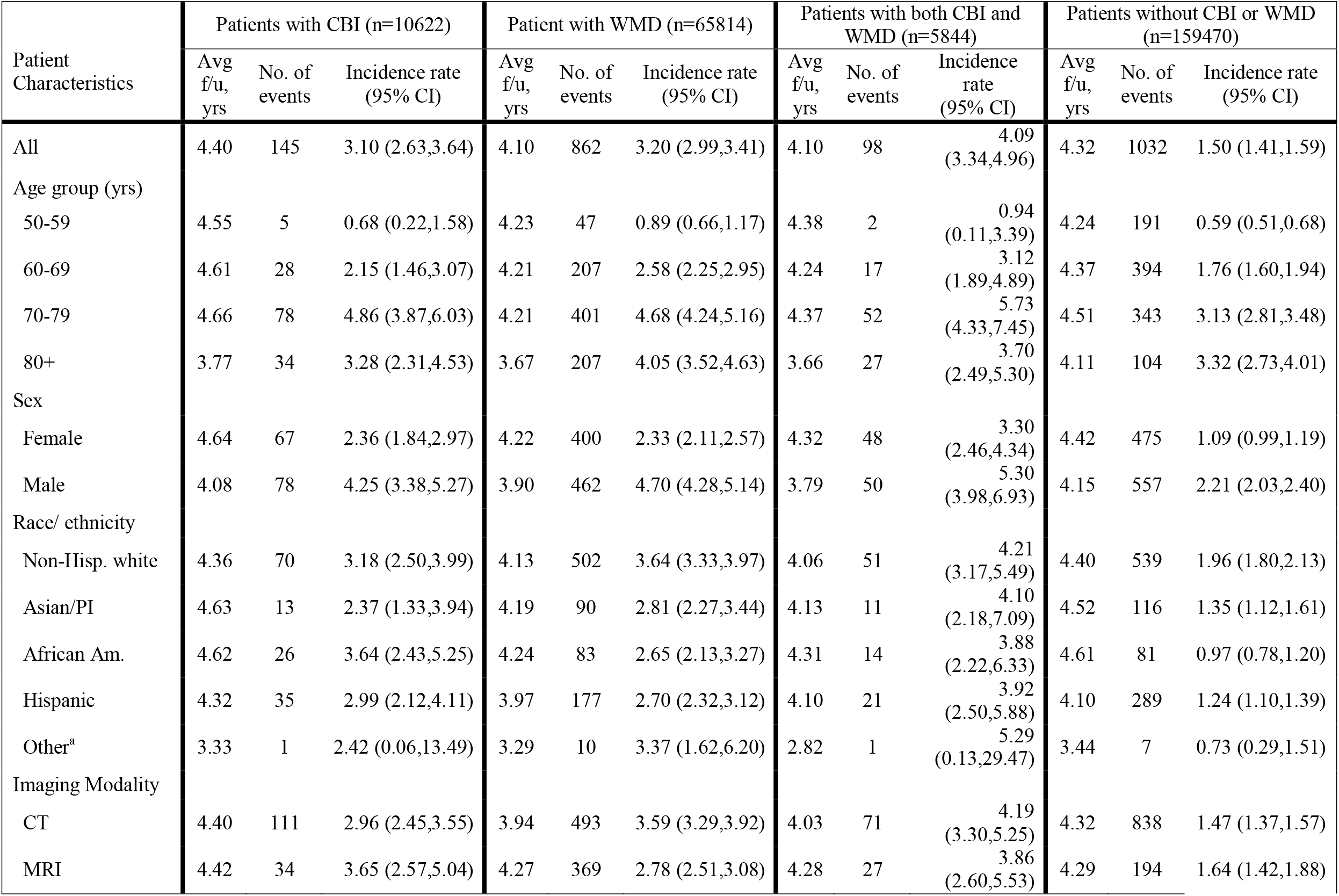
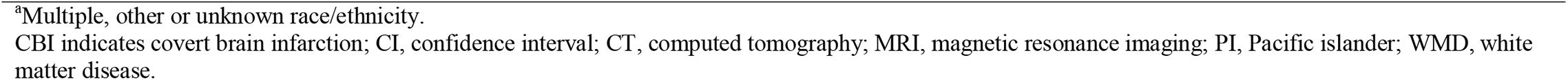
Parkinson’s disease incidence rate overall and in subgroups (n=230,062).

| Patient Characteristics | Patients with CBI (n=10622) |  |  | Patient with WMD (n=65814) |  |  | Patients with both CBI and WMD (n=5844) |  |  | Patients without CBI or WMD (n=159470) |  |  |
| --- | --- | --- | --- | --- | --- | --- | --- | --- | --- | --- | --- | --- |
|  | Avg f/u, yrs | No. of events | Incidence rate (95% CI) | Avg f/u, yrs | No. of events | Incidence rate (95% CI) | Avg f/u, yrs | No. of events | Incidence rate (95% CI) | Avg f/u, yrs | No. of events | Incidence rate (95% CI) |
| All | 4.40 | 145 | 3.10 (2.63,3.64) | 4.10 | 862 | 3.20 (2.99,3.41) | 4.10 | 98 | 4.09 (3.34,4.96) | 4.32 | 1032 | 1.50 (1.41,1.59) |
| Age group (yrs) |  |  |  |  |  |  |  |  |  |  |  |  |
| 50-59 | 4.55 | 5 | 0.68 (0.22,1.58) | 4.23 | 47 | 0.89 (0.66,1.17) | 4.38 | 2 | 0.94 (0.11,3.39) | 4.24 | 191 | 0.59 (0.51,0.68) |
| 60-69 | 4.61 | 28 | 2.15 (1.46,3.07) | 4.21 | 207 | 2.58 (2.25,2.95) | 4.24 | 17 | 3.12 (1.89,4.89) | 4.37 | 394 | 1.76 (1.60,1.94) |
| 70-79 | 4.66 | 78 | 4.86 (3.87,6.03) | 4.21 | 401 | 4.68 (4.24,5.16) | 4.37 | 52 | 5.73 (4.33,7.45) | 4.51 | 343 | 3.13 (2.81,3.48) |
| 80+ | 3.77 | 34 | 3.28 (2.31,4.53) | 3.67 | 207 | 4.05 (3.52,4.63) | 3.66 | 27 | 3.70 (2.49,5.30) | 4.11 | 104 | 3.32 (2.73,4.01) |
| Sex |  |  |  |  |  |  |  |  |  |  |  |  |
| Female | 4.64 | 67 | 2.36 (1.84,2.97) | 4.22 | 400 | 2.33 (2.11,2.57) | 4.32 | 48 | 3.30 (2.46,4.34) | 4.42 | 475 | 1.09 (0.99,1.19) |
| Male | 4.08 | 78 | 4.25 (3.38,5.27) | 3.90 | 462 | 4.70 (4.28,5.14) | 3.79 | 50 | 5.30 (3.98,6.93) | 4.15 | 557 | 2.21 (2.03,2.40) |
| Race/ ethnicity |  |  |  |  |  |  |  |  |  |  |  |  |
| Non-Hisp. white | 4.36 | 70 | 3.18 (2.50,3.99) | 4.13 | 502 | 3.64 (3.33,3.97) | 4.06 | 51 | 4.21 (3.17,5.49) | 4.40 | 539 | 1.96 (1.80,2.13) |
| Asian/PI | 4.63 | 13 | 2.37 (1.33,3.94) | 4.19 | 90 | 2.81 (2.27,3.44) | 4.13 | 11 | 4.10 (2.18,7.09) | 4.52 | 116 | 1.35 (1.12,1.61) |
| African Am. | 4.62 | 26 | 3.64 (2.43,5.25) | 4.24 | 83 | 2.65 (2.13,3.27) | 4.31 | 14 | 3.88 (2.22,6.33) | 4.61 | 81 | 0.97 (0.78,1.20) |
| Hispanic | 4.32 | 35 | 2.99 (2.12,4.11) | 3.97 | 177 | 2.70 (2.32,3.12) | 4.10 | 21 | 3.92 (2.50,5.88) | 4.10 | 289 | 1.24 (1.10,1.39) |
| Other<sup>a</sup> | 3.33 | 1 | 2.42 (0.06,13.49) | 3.29 | 10 | 3.37 (1.62,6.20) | 2.82 | 1 | 5.29 (0.13,29.47) | 3.44 | 7 | 0.73 (0.29,1.51) |
| Imaging Modality |  |  |  |  |  |  |  |  |  |  |  |  |
| CT | 4.40 | 111 | 2.96 (2.45,3.55) | 3.94 | 493 | 3.59 (3.29,3.92) | 4.03 | 71 | 4.19 (3.30,5.25) | 4.32 | 838 | 1.47 (1.37,1.57) |
| MRI | 4.42 | 34 | 3.65 (2.57,5.04) | 4.27 | 369 | 2.78 (2.51,3.08) | 4.28 | 27 | 3.86 (2.60,5.53) | 4.29 | 194 | 1.64 (1.42,1.88) |

<sup>a</sup>Multiple, other or unknown race/ethnicity.
CBI indicates covert brain infarction; CI, confidence interval; CT, computed tomography; MRI, magnetic resonance imaging; PI, Pacific islander; WMD, white matter disease.

### Follow-up

Follow-up started 180 days after the index scan and ended with the earliest of the following events: disenrollment from the health plan, end of the study (December 31, 2019), death, Parkinson’s disease (outcome), or other censoring events of stroke and dementia.

### Outcome definition

The primary outcome of this study was Parkinson’s disease, defined by the following codes: ICD-9 = 332.0 and ICD-10 = G20. In our primary analysis, we required 2 diagnoses, with the time of onset at the first diagnosis.

### Statistical Analysis

Kaplan-Meier plots were used to present PD-free survival in patients with and without CBI and in patients with and without WMD. The differences in the distributions between patients with and without CBI (WMD) were assessed by the log-rank test. The overall and risk-factor stratified crude incidence rates and the 95% confidence intervals (CI) were calculated using Poisson regression models and reported as per 1,000 person-years of follow-up time. We examined the crude and adjusted associations of CBI and WMD with PD, using Cox proportional hazards regression models. Multiple imputation was used to account for missing data under the missing at random assumption, and estimates were pool across ten imputed datasets based on Robin’s rule.^17,18^ For adjusted effects, we included known cardiovascular risk factors for stroke based on prediction models in the literature, including the following covariates: age group, sex, race/ethnicity, atrial fibrillation, carotid atherosclerosis, congestive heart failure, coronary artery disease, diabetes mellitus, hypercholesterolemia, hypertension, peripheral arterial disease, ever tobacco use, mean systolic blood pressure in one year, antiplatelet use, stain use, depression, exercise (hour/per week), body mass index (BMI, kg/m^2^), and modality.^19,20^ Systolic blood pressure, exercise and BMI were imputed using multiple imputation^17^ when they were missing (3.68%, 14.83% and 1.00%, respectively).

To test the proportional hazards assumption, we examined the independence of the Schoenfeld residuals and follow-up time for each variable. Interaction terms were selected based on clinical judgment. We hypothesized that the effects of WMD and CBI would vary based on imaging modality. We thus compared the effect of the presence versus the absence of CBI or WMD separately in those examined by CT and by MRI. We also anticipated that these lesions might have greater prognostic importance in younger versus older patients and so included interactions with age (< age 70 versus ≥ age 70).

To examine the effects of WMD severity, analyses were performed similarly to the main analysis described above. However, WMD severity was interacted with imaging modality and thus treated as 10 different classes (i.e., no WMD, mild WMD, moderate WMD, severe WMD, and undetermined WMD for CT and for MRI, respectively). The reference class for all hazards was those patients who underwent MRI imaging and were found to be free of WMD (i.e., the lowest risk group). We underscore that this approach, using a common reference class, is distinct from the main analysis which contrasted PD hazards for patients with and without CCD for each modality separately (e.g., comparing hazards for PD among patients with versus without WMD among those undergoing CT and separately among those undergoing MRI, using different reference classes for these contrasts).

Analyses were performed using SAS (Version 9.4 for Unix; SAS Institute, Cary, NC) and R Version 3.6.0 (R Foundation, Vienna, Austria).

### Sensitivity Analysis

Because symptoms of PD may precede the diagnosis of PD substantially and may also prompt neuroimaging, we performed a sensitivity analysis that excluded patients with a diagnosis of PD within one year of the index scan. For this analysis, “time zero” was one year after the index scan. We also performed sensitivity analyses using two more specific definitions of PD: 1) requiring at least one of the two PD diagnoses to be made by a neurologist; 2) requiring at least two prescriptions of a single anti-PD medication. The purpose of this was to examine the effects of more carefully excluding vascular parkinsonism and other secondary forms of parkinsonism.

## RESULTS

A total of 230,062 patients meeting inclusion criteria were included, with a total of 980,772 person-years of follow-up time (Supplement B, eFigure 1). The median follow-up time was 3.72 years (range of 181 days to 10.50 years; IQR 1.52 to 6.80 years). There were 1,941 cases of PD identified in follow-up, with a median time-to-event of 2.35 (IQR 0.90 to 4.58) years. Table 1 describes patient characteristics in the total cohort and in those with CBI (regardless of WMD) and those with WMD (regardless of CBI). 59124 (25.70%) patients underwent MRI scans and 170938 (74.30%) underwent CT scans. CCD was identified in 70592 (30.68%) patients, 10622 (4.62%) with CBI and 65814 (28.61%) with WMD shown in Table 2. The crude PD incidence rates (per 1000 person-years) were 3.10 (2.63, 3.64) in patients with incidentally discovered CBI and 3.20 (2.99, 3.41) for patients with WMD. For patients with both CBI and WMD, the incidence rate was 4.09 (3.34, 4.96). This compares to an incidence rate of only 1.50 (1.41, 1.59) in patients free of cerebrovascular disease. The crude hazard ratio (HR) for PD associated with WMD was 2.10 (1.92, 2.29); and with CBI was 1.62 (1.36, 1.91), as shown in Table 3.

**Figure 1.**
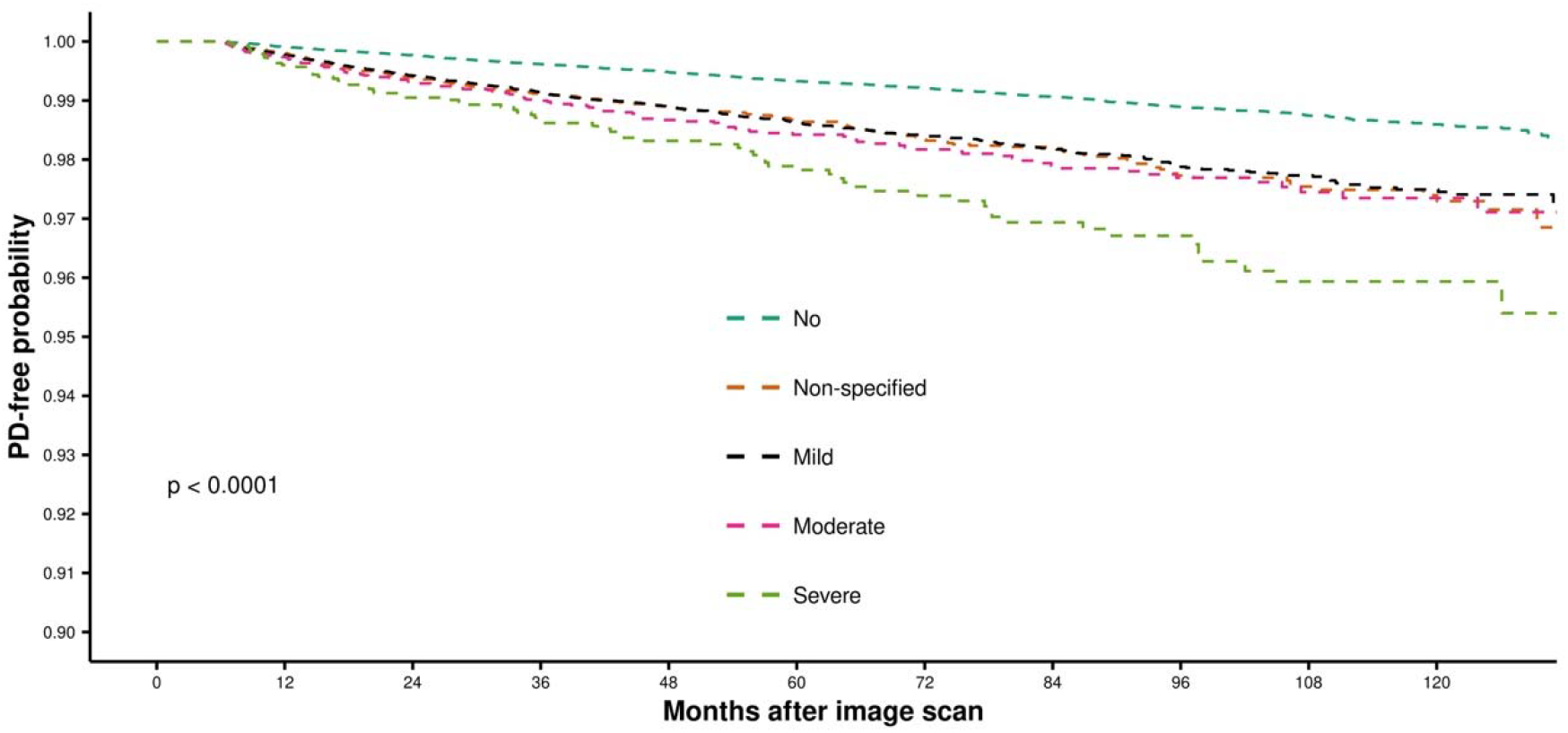
Kaplan-Meier curves depicting stroke-free survival by white matter disease severity grade.

**Table 3.**
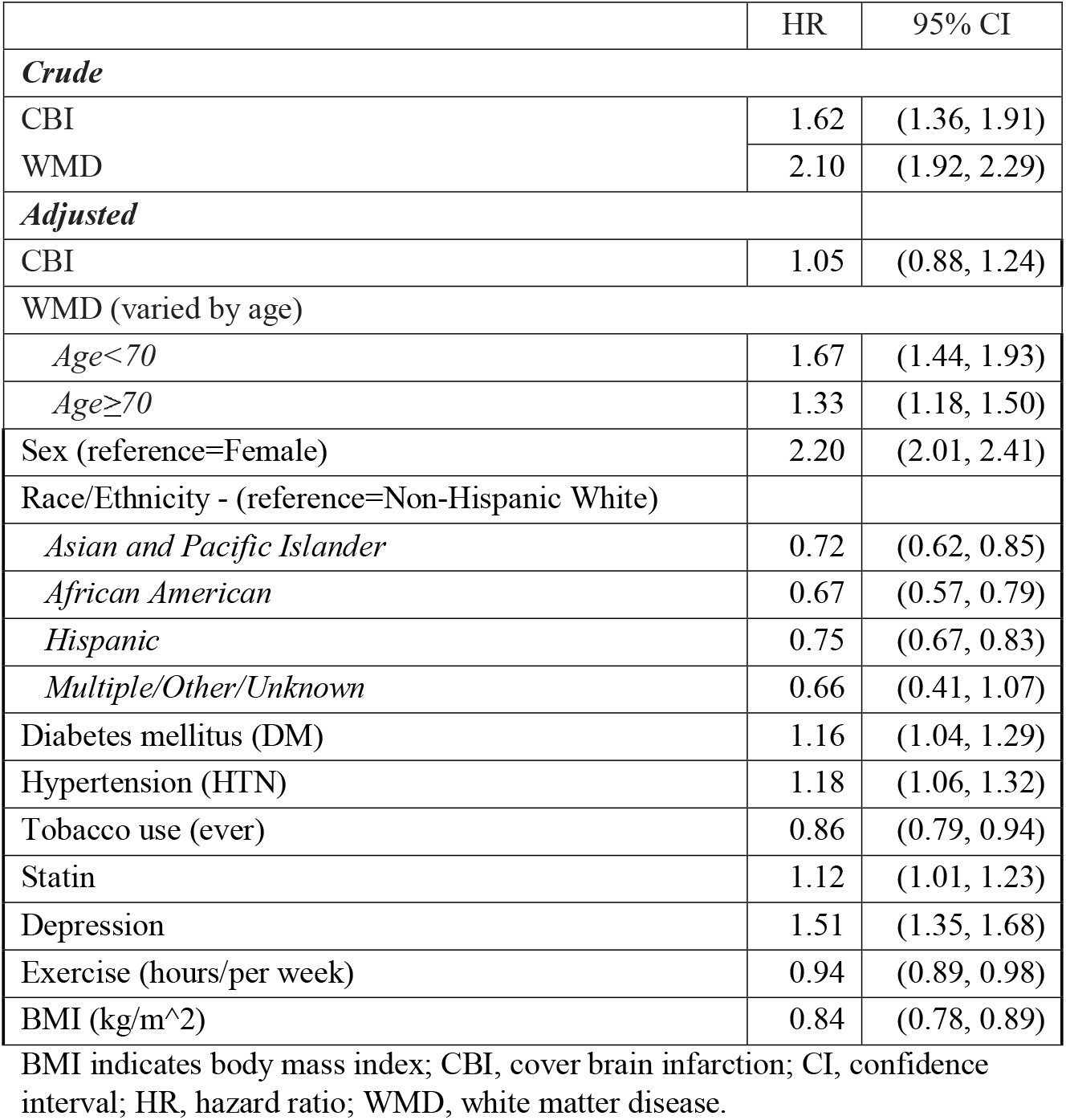
Crude and adjusted hazard ratios (HRs) for Parkinson’s disease by covert brain infarction (CBI) and white matter disease (WMD).

|  | HR | 95% CI |
| --- | --- | --- |
| *Crude* |  |  |
| CBI | 1.62 | (1.36, 1.91) |
| WMD | 2.10 | (1.92, 2.29) |
| *Adjusted* |  |  |
| CBI | 1.05 | (0.88, 1.24) |
| WMD (varied by age) |  |  |
| *Age<70* | 1.67 | (1.44, 1.93) |
| *Age≥70* | 1.33 | (1.18, 1.50) |
| Sex (reference=Female) | 2.20 | (2.01, 2.41) |
| Race/Ethnicity - (reference=Non-Hispanic White) |  |  |
| *Asian and Pacific Islander* | 0.72 | (0.62, 0.85) |
| *African American* | 0.67 | (0.57, 0.79) |
| *Hispanic* | 0.75 | (0.67, 0.83) |
| *Multiple/Other/Unknown* | 0.66 | (0.41, 1.07) |
| Diabetes mellitus (DM) | 1.16 | (1.04, 1.29) |
| Hypertension (HTN) | 1.18 | (1.06, 1.32) |
| Tobacco use (ever) | 0.86 | (0.79, 0.94) |
| Statin | 1.12 | (1.01, 1.23) |
| Depression | 1.51 | (1.35, 1.68) |
| Exercise (hours/per week) | 0.94 | (0.89, 0.98) |
| BMI (kg/m<sup>2</sup>) | 0.84 | (0.78, 0.89) |

BMI indicates body mass index; CBI, covert brain infarction; CI, confidence interval; HR, hazard ratio; WMD, white matter disease.

In a multivariable model controlling for major cardiovascular risk factors and other factors known to be associated with PD, the presence of CBI was no longer found to be significantly associated with PD (HR=1.05 [0.88, 1.24]) (Table 3). The effect of WMD was found to vary by age. For patients younger than age 70 the adjusted HR was 1.67 (1.44, 1.93) and for that age 70 or older the HR was 1.33 (1.18, 1.50). Effects were consistent across imaging modalities. The effects of other risk factors on PD incidence were generally consistent with the literature^21,22^, as shown in Table 3. PD incidence was associated with male, non-white ethnicity, cardiovascular risk factors (e.g., diabetes and hypertension), and the use of statin medication. PD was also strongly associated with depression. Smoking, exercise, and increased BMI were associated with a decreased risk of PD.

### Analysis of WMD grade

As shown in Table 4, the incidence of PD (per 1,000 person-years) was found to increase with increasing severity of WMD, from 1.52 (1.43, 1.61) for patients without WMD to 4.90 (3.86, 6.13) for those with severe WMD. The adjusted hazard ratio was 1.88 (1.47, 2.41) for those with severe WMD.

**Table 4.** Incidence rate and crude and adjusted hazard ratios (HR) of white matter disease (WMD) and risk of Parkinson’s disease by severity (n=230,062).

| WMD Severity | Number of patients | Average f/u, years | Number of events | Incidence rate/1,000 person years (95% CI) | Crude HR (95% CI) | Adjusted HR* (95% CI) |
| --- | --- | --- | --- | --- | --- | --- |
| none (ref) | 164248 | 4.33 | 1079 | 1.52 (1.43, 1.61) | - | - |
| mild | 38548 | 4.10 | 477 | 3.02 (2.76, 3.30) | 1.98 (1.78,2.21) | 1.40 (1.25,1.58) |
| moderate | 8643 | 4.13 | 121 | 3.39 (2.83, 4.04) | 2.23 (1.85,2.69) | 1.38 (1.13,1.69) |
| severe | 3462 | 4.25 | 72 | 4.90 (3.86, 6.13) | 3.22 (2.54,4.09) | 1.88 (1.47,2.41) |
| not specified | 15161 | 4.05 | 192 | 3.13 (2.71, 3.60) | 2.05 (1.76,2.39) | 1.32 (1.13,1.55) |

\*Adjusting for age group, sex, race/ethnicity, atrial fibrillation, carotid atherosclerosis, congestive heart failure, coronary artery disease, diabetes mellitus, hypercholesterolemia, hypertension, peripheral arterial disease, ever tobacco use, mean systolic blood pressure in one year, antiplatelet use, statin use, depression, exercise (hour/per week), BMI (kg/m<sup>2</sup>), modality, and CBI.
CI indicates confidence interval; HR, hazard ratio; WMD, white matter disease

Kaplan-Meier curves demonstrating PD-free survival probability of patients with WMD stratified by severity are shown in Figure 1. Increasing severity of WMD was associated with lower PD-free probability.

### Sensitivity Analyses

When excluding patients on antithrombotic medications/indications, a diagnosis of PD within one year of the index scan, or when using more specific definitions of PD (i.e., requiring neurologist diagnosis or anti-PD medications), the incidence rates decreased somewhat, but crude and adjusted hazard ratios for CBI and for WMD remained stable (Supplement B, eTables 1 to 8). For example, when only including patients with at least one year of follow up, the crude HR for PD associated with WMD was 2.04 (95% CI 1.38 to 1.99) and the adjusted HR was 1.59 (95% CI 1.34 to 1.87) in patients younger than 70 and 1.29 (95% CI 1.13 to 1.47) in older patients.

## DISCUSSION

In the present study, we examine the association of incidentally discovered CCD with PD, leveraging NLP of neuroimaging reports obtained during routine clinical care. WMD was found to be associated with subsequent PD. Stratification by age, revealed that the association was stronger in younger individuals (adjusted HR for age less than 70 is 1.67 [1.44, 1.93] and for age 70 or older is 1.33 [1.18, 1.50]). The reason for this difference is not immediately apparent, but might include the greater difficulty diagnosing PD againsts a background of diminished age-related activity and competing events in older persons, as well as an age-dependent increase in PD risk from pathways unrelated to cerebrovascular disease. Furthermore, we observed that the risk of PD rose with the increasing severity of WMD, and this ‘dose-dependency’ adds credence to the overall association. Conversely, CBI was not found to be associated with subsequent PD after adjusting for other cardiovascular and PD risk factors.

Understanding the risk factors that predispose individuals to PD can potentially inform prevention strategies, both through identifying vulnerable patients (i.e., predictive inference) and identifying potential therapeutic targets (i.e., causal inference). To help identify patients at-risk of developing PD, work has begun developing predictive algorithms using combinations of early features of PD and risk factors, though they do not currently consider CCD as one of the predictive factors.^23-25^ The magnitude of the effect with WMD is substantially larger than that typically observed for many individual cardiovascular risk factors and similar to depression.^21,22^ It is also similar in magnitude to the effect of prior stroke reported in previous studies.^4,26^ Incorporating CCD in risk prediction in clinical care however is limited by the absence of neuroimaging results in many patients at risk for PD.

We are aware of only one prior cohort study to examine the relationship between small vessel disease, including WMD, and the risk of incident parkinsonism^3^. This study included 501 patients, only 20 only 20 of whom developed the outcome. While a positive association was observed, this was due largely to patients with vascular parkinsonism, which accounted for 15 of the 20 outcomes. In contrast, our study included almost 2000 patients who received an incident diagnosis of PD. It is the first cohort study we are aware of to find an association between WMD and a subsequent diagnosis of PD. While some of these cases may have been misdiagnosed vascular parkinsonism, PD is the most common cause of parkinsonism and our results were robust even when the diagnosis was restricted to more specific outcome definitions (including requiring a neurologists diagnosis and requiring sustained anti-PD medications).

PD is characterized by degeneration of dopaminergic neurons in the substantia nigra and hallmark Lewy pathology in surviving neurons. Our findings raise the possibility that cerebrovascular disease including covert WMD might promote PD pathology, given prior studies evidence, the large size of the effect, the monotonic ‘dose response’ relationship with WMD severity, and the temporality. While idiopathic PD has traditionally been thought to arise from toxic-metabolic injury, our results suggest that cerebrovascular injury may also play a role in promoting or facilitating neurodegeneration. Putative mechanism include ischemia-induced inflammation, neuroglial activation, and subsequent neurodegeneration.^27-29^

We note that prior case-control and pathological studies have found an inconsistent association between cerebrovascular disease and PD.^12^ These cross-sectional studies are more prone to issues of reverse causation than cohort studies, since cohort studies can select patients free of PD at baseline and follow them over time. Prior investigators have speculated that PD may reduce the risk of cerebrovascular disease, either through lower levels of dopamine in the brain or as a result of the blood pressure lowering effects of levodopa, which might obscure the relationship between prior cerebrovascular disease and PD.^30-32^ Our approach using NLP and routinely obtained neuroimaging reports permitted us to leverage a very large database obviating the need for, and overcoming the limitations of, a case-control approach.

For years, discussion of the role of vascular risk factors and PD was dominated by the well-known inverse association between smoking and risk of PD.^33^ Whether or not lower risk of PD is causally driven by smoking in cigarette smoke (such as nicotine) or the association is explained by reverse causation, or another bias (such as survival bias), remains unclear. It is striking that in observational studies prone to survival bias, inverse associations have been documented for many vascular risk factors, presumably because the mortality associated with these can occur before PD is ascertained as an outcome.^33^ However, in recent times the relationship between other vascular risk factors and PD has been under more scrutiny. Most notable is an increasingly robust association between type 2 diabetes (T2D) and risk of PD.^34^ Support from epidemiology and the investigation of overlapping mechanisms between T2D and PD mean that repurposed drugs used to treat T2D are among those considered most likely to yield disease-modifying opportunities for PD.^35^ Higher BMI is a challenge to study, because BMI is known to change early in the clinical phases of PD. Higher BMI in a meta-analysis of prospective observational studies was not found to be associated with PD^36^ but a null association has not always been observed^37^, genetically estimated higher BMI has been associated with lower risk of PD in Mendelian randomization studies^38^.

Hypertension too, while generally showing an inverse association with PD in retrospective case-control studies, has been shown to be associated with future PD in some settings^39^. The relationship between cholesterol and other lipids and PD remains unclear. Combining many of these factors into a concept of a metabolic syndrome has been done and suggests that metabolic syndrome and its components may be risk factors for PD.^40^ Finally, a Mediterranean diet is renowned for reducing cardiovascular risk, and has also been linked to lower risk of PD.^41,42^

There were several limitations to this study. First, we rely on diagnostic codes from routine clinical care. This may result in misclassification which often attenuate associations. Similarly, some patients classified as having idiopathic PD may have had vascular parkinsonism from basal ganglia infarcts or severe periventricular white matter disease, which may lead to a spurious association. Nevertheless, more stringent definitions of the PD outcome did not substantially attenuate the observed associations, including those requiring sustained anti-PD medication. Second, we relied on neuroimaging obtained through routine care, which is obtained only when clinically indicated and is therefore subject to selection bias, which can potentially distort causal associations. Yet for prediction, this cohort is arguably the clinically relevant target, as these patients have come to attention through routine care. Furthermore, relying on NLP of neuroimaging reports may be criticized for consistency and reliability compared to advanced imaging analysis-based approaches (e.g., volumetric). However, this may also be viewed as a strength as these are the types of reports that come to attention during everyday clinical practice. Our study utilizes a large, real-world, population-level cohort that overcomes the limitations of smaller studies. Using real-world data also permits insights not afforded from research cohorts— particularly the evaluation of real-world effects, including associations of findings discovered by both CT scan and MRI among patients with clinically indicated scans and an understanding of the prognostic information contained in routinely obtained neuroimaging reports. Finally, observational studies such as this do not provide evidence of causation without unverifiable assumptions; this study shows that WMD identified on routinely obtained neuroimages identifies patients at higher risk of developing PD. The causal role of these lesions in PD development would be strengthend by intervention studies targeting cerebral ischemia or infarction.

In summary, we found that WMD identified by NLP of routinely obtained neuroimages was associated with subsequent a PD diagnosis, an association strengthened with younger age and increased WMD severity, and unaffected by more stringent outcome definitions. CBI did not seem to be associated with subsequent PD when adjusted for risk factors.

## Data Availability

All data produced in the present work are contained in the manuscript.

## Acknowledgements

Dr. Kent had full access to all the data in the study and takes responsibility for the integrity of the data and the accuracy of the data analysis.

This work was funded by a National Institutes of Health (NIH) grant (R01-NS102233). The funder had no role in the design and conduct of the study; collection, management, analysis, and interpretation of the data; preparation, review, or approval of the manuscript; and decision to submit the manuscript for publication.

## Author Contributions

DMK contributed to conceptualization, funding acquisition, investigation, methodology, supervision, and wrote the original manuscript draft. LYL, EJP, PHL, DFK, and SF contributed to investigation and methodology. AYW and AJN contributed to drafting a significant portion of the manuscript draft. JN contributed to investigation. CZ contributed to data curation, investigation, and methodology. HL contributed to funding acquisition, investigation, methodology, and supervision. WC contributed to data curation, formal analysis, investigation, methodology, and supervision. All authors contributed equally to reviewing and editing the final manuscript.

